# Management implications of shifting *Culex quinquefasciatus* West Nile virus transmission suitability in Florida

**DOI:** 10.1101/2025.09.22.25336394

**Authors:** Sadie J. Ryan, Catherine A. Lippi

## Abstract

**Background:** West Nile virus (WNV), a mosquito-borne flavi*virus*, has circulated in the USA since 1999. In 2025, Florida was home to 24 million people, with projected increases in population and urbanization in a changing climate. The southern house mosquito, *Culex quinquefasciatus*, is found in every county, and is a major vector for WNV. Describing shifting WNV transmission risk is important to inform public health and vector control planning.

**Materials and Methods:** Using published estimates of thermal suitability of WNV transmission by *Cx quinquefasciatus*, with climate models and population data, we calculated and mapped baseline and projected county-level transmission suitability and people at risk (PAR) for 2000, 2030, and 2050. Five general circulation models (GCMs) and two mitigation scenarios (SSP2-4.5, SSP5-8.5) were used to explore future trajectories.

**Results:** At baseline, all 67 counties in Florida experienced 5-9 months transmission suitability. Using the year 2000 census estimates, 2.33 million people in 2 counties experienced 9 months, and in 2030, across climate models, 8.93-12.26 million people (10-20 counties; SSP2-4.5), and 8.95-18.10 million people (11-26 counties; SSP5-8.5) are projected to experience 9 or more months of transmission suitability. In 2050, for both SSP2-4.5 and SSP5-8.5, 17.08-20.42 million people (23-26 counties), ranging to approximately 70% of the projected population of Florida will experience 9 or more months. The 10 most populated counties in 2000 are projected to experience 1-3 months of additional climate driven transmission suitability in the future.

**Conclusion:** The southern house mosquito was previously managed as a seasonal nuisance in Florida but now represents an increasing public health exposure risk. Projections across climate trajectories underscore an increasing suitability and exposure risk for WNV in Florida, ranging as high as around 70% of the population exposed to suitable climate conditions for transmission for 9 or more months of the year in the 2050s. This means the types of operations and number of employees needed in vector control and public health will also increase.

## Introduction

Geospatial patterns of mosquito-borne diseases are anticipated to shift in response to global change processes, including global warming and rapidly urbanizing landscapes with growing populations (Bhattarai et al., 2022; Ryan, 2020; Ryan et al., 2023, 2019). The thermal dependency of the tightly coupled relationship between mosquitoes and pathogens, and their roles in transmission, have been explored for several mosquito-borne diseases typically thought of as tropical (Miazgowicz et al., 2020; Mordecai et al., 2019, 2017, 2013; Shocket et al., 2018; Tesla et al., 2018), and also for several temperate flaviviral diseases (Mordecai et al., 2019; Shocket et al., 2020). These temperature dependent transmission relationships have been leveraged in previous work to explore the geographic bounds of transmission suitability, and the corresponding seasonal suitability of transmission, to translate non-linear temperature dependent transmission functions into a mapped risk estimate (Miazgowicz et al., 2020; Tesla et al., 2018). This has been used to assess areas suitable for transmission, and to quantify the number of people at risk (PAR) of potential transmission. By extending this approach via coupling transmission suitability to climate projections, the potential for increases and decreases in the number of people at risk, in addition to the geographic redistribution of this risk, under future scenarios can be quantified and visualized (Rohat et al., 2020; Ryan, 2020; Ryan et al., 2021, 2020, 2019, 2015). This creates essential tools to plan, anticipate, and even target strategies for intervention and mitigation (Ryan et al., 2020).

West Nile virus (WNV), an introduced arboviral disease in North America, is widely established (Kramer et al., 2019). Manifesting in periodic outbreaks since its first North American detection in 1999, WNV is transmitted to humans as part of a spillover from a sylvatic cycle (Eidson et al., 2001; Nash et al., 2001). It is well suited to reservoirs of birds, particularly corvids and other passerines, and the disease often becomes apparent in birds when they are found sick or dead by people (Eidson et al., 2001). Humans are infected when bitten by the same *Culex* spp. mosquitoes that bite the birds (Kilpatrick et al., 2005), and are considered a dead-end host (Colpitts et al., 2012). Similarly to multiple other flaviviral infections in humans, a large proportion of cases are subclinical or asymptomatic, and thus go unreported and largely undocumented. Symptomatic cases manifest as a range from fever and myalgias, to becoming neuroinvasive and leading to meningoencephalitis, and even death (Colpitts et al., 2012; Petersen and Marfin, 2002). Therefore, reported human cases represent a small fraction of potential human cases, with the majority of reported cases in the USA being neuroinvasive.

Neuroinvasive WNV appears to be most severe in older individuals, with case fatality rates of 15-29% reported in 70+ year-olds (Petersen and Marfin, 2002), and the sequelae (onward impact) of neurological implications for cases of all ages can be significant. While native species of birds have been implicated, the domestic bird reservoir for WNV in North America comprises a suite of birds that were introduced over the past several hundred years. Thus, the continued cycling of WNV is likely only constrained by the range of its mosquito vectors, primarily *Culex* spp. In Florida, human WNV cases are reported every year (average 7 non-neuroinvasive, 18 neuroinvasive), with 501 (390 neuroinvasive), human cases reported to the CDC from 1999-2024 (CDC, 2025). As these data indicate, neuroinvasive cases are more severe, and thus more likely to be reported – but non-neuroinvasive cases are also sufficiently severe to lead to reportable clinical diagnoses. In 2020, at the height of COVID-19 related lockdowns and travel restrictions, there was ongoing WNV transmission in Florida, reflected in 51 reported human cases and viral presence detected in mosquitoes (Coatsworth et al., 2022). In 2025, Alachua County in North Central Florida released public health alerts when WNV was detected in sentinel chickens (DOH-Alachua, 2025). In Florida, WNV is an ongoing and present flaviviral disease risk, for which case surveillance is complicated by clinical manifestation, but with potential for severe public health impacts.

In previous work, Shocket et al (2020) established the thermal boundaries of transmission of WNV by *Culex quinquefasciatus*, whose range in the USA spans from Florida to southern California, an expansive distribution, only potentially constrained by temperature bounds. Work by Wilke et al (Wilke et al., 2021) demonstrated that in Florida, urban environments create an ecological filtering on the community of mosquito species, favoring *Aedes aegypti* and *Cx. quinquefaciatus*. This points to the increasing risk of WNV transmission in an increasingly populated and urbanizing environment. Florida is not unique in the USA in its patterns of increasing urbanization due to population increases, but it is among the most populous states in the USA, ranked 3^rd^ after California and Texas <u>(US Census Bureau, 2026)</u>. In coming years,

Florida is predicted to continue its population increase and increase in population density patterns (Rayer and Wang, 2021), and existing population centers will continue to both expand and infill, creating a dual pattern of increased numbers of people, and increased urbanization of the landscape and density of people. While these may generate similar implications for epidemiological risk at a state level, WNV, essentially specializing on Florida urbanizing environments, presents both amplified, and geographically specific shifts in risk in coming years.

In this study, we quantify and map thermal suitability for WNV transmission in months, and an estimate of PAR, at the county level, in Florida, at baseline climate conditions, and for 2021-2040 (denoted 2030) and 2041-2060 (denoted 2050) projected climate under two potential mitigation trajectories, and five CMIP6 general circulation models (GCMs - see Methods for details). The two trajectories represent a mitigation (SSP2-4.5) and no mitigation (SSP5-8.5) scenario, enabling us to examine the geographic and demographic shifts in risk of WNV transmission by *Cx. quinquefasciatus* anticipated within the decade (2030s), and in a generation (2050s). While additional factors comprise the interaction of humans with the potential transmission of WNV, changing temperature and the geographic pattern of projected population, are key to understanding and anticipating changing risk and potential management strategy needs. In this study, we quantify projected changes for Florida at the county and state levels, and for illustration, use the ten currently most populated counties, spanning a North-South gradient, (i.e., Duval, Volusia, Orange, Polk, Pinellas, Hillsborough, Brevard, Palm Beach, Broward, Miami-Dade) to showcase WNV transmission suitability across climate models, scenarios, and years. Vector management, including the presence and staffing of vector control districts, allocation of budgetary resources to surveillance activities, and even availability of equipment, capacity, and type of response, differs by county across the state of Florida.

Understanding the potential for future needs via quantified changes in season, geography, and populations at risk, can help inform planning needs across the state. Hiring seasonal vector control technicians may evolve into hiring more permanent staff with these projected shifts in risk duration and magnitude. To make our findings accessible and usable, we provide our raster-based model output and a general code pipeline as open, publicly available data (https://github.com/sjryan3/wnv_FL) for reuse and re-mapping for decision making needs.

## Materials and Methods

### WNV Thermal Transmission Suitability

Using a published experimentally derived mechanistic model of WNV transmission by *Cx. quinquefasciatus* (Shocket et al., 2020), we calculated transmission suitability as a function of temperature (*S(t)*) for Florida. The thermal limits for WNV transmitted by *Cx. quinquefasciatus* are 19.0 °C (14.1-20.9) and 31.8 °C (31.1-32.3), with optimal transmission occurring at 25.2 °C (23.9-27.1) (Shocket et al., 2020). A similar methodological process as prior mapping studies for thermal suitability shifts in a changing climate was followed (e.g. (Ryan et al., 2023, 2019)). To capture a conservative estimate of the thermal bounds, we mapped *S(t)* at the lower 95% posterior credibility interval.

### Climate Data

Mean monthly temperatures were obtained from WorldClim.org. Monthly mean temperature rasters at 30s resolution (approximately 1km^2^ at the equator) were downloaded for baseline (1970-2000), and monthly minimum (tmin) and maximum temperatures (tmax) for projected scenarios (2021-2040, ‘2030’) and (2041-2060, ‘2050’), using WorldClim version 2.1 CMIP6 data products. Scenarios corresponding to representative concentration pathways RCP 4.5 and 8.5 were chosen, described as SSP2-4.5 and SSP5-8.5 in CMIP6 scenarios, to compare potential climate change outcomes as a function of projected mitigation levels.

To account for an issue in CMIP6 models used for AR6 IPCC reporting, known colloquially as “the hot model issue”, wherein several general circulation models (GCMs) included in the AR6 ensemble predicted larger temperature increases, and may have introduced a hotter bias into interpreted description of impact (Hausfather et al., 2022), we chose to use separate GCMs for our projected climate scenarios as follows. We chose to follow suggestions to examine two key sensitivity metrics, transient climate response, TCR, and equilibrium climate sensitivity, ECS, and choose GCMs spanning a range of sensitivity. Hausfather et al <u>(2022)</u> describe their usage of these, where TCR = 1.4-2.2°C falls in the ‘likely’ range, and ECS=2.5-4°C falls in the ‘likely’ range. Scaffetta <u>(2023)</u> examined these further, assigning groupings of ECS and TCR ranges into low (L), medium (M), and high (H) ranges, in which the low sensitivity group best reproduced existing warming in their study. We thus assigned a ranking to five GCMs, spanning these sensitivity ranges, according to Scafetta’s groupings using values of TCR and ECS to designate H, M, L for each sensitivity metric and create a ranking for this study from “hottest” to “coldest” projection model (Table 1).

**Table 1.** The five general circulation models (GCMs) selected. (abbreviations and full model names) to illustrate the range of model outcomes within the CMIP6 suite, using measures of model sensitivity (transient climate response, TCR, and equilibrium climate sensitivity, ECS), designated Low (L), Medium (M) and High (H) to rank ‘hottest’ to ‘coldest’ projections (see methods).

| Abb | GCM | ECS (°C) | TCR (°C) | Ranking |
| --- | --- | --- | --- | --- |
| HAD | HadGEM3-GC31-LL | 5.55 | 2.55 | HH (hottest) |
| ACC | ACCESS-CM2 | 4.72 | 2.10 | HM |
| CMC | CMCC-ESM2 | 3.55 | 1.92 | MM |
| MRI | MRI-ESM2-0 | 3.15 | 1.64 | ML |
| MIR | MIROC6 | 2.61 | 1.55 | LL (coldest) |

### Demographic Data

County level census data for Florida from the 2000 US Census were used to align to the baseline climate scenario (U.S. Census Bureau, 2017). County-level projected population estimates for 2030 and 2050 were obtained from the Florida Population Studies Program (Rayer and Wang, 2021).

### Analyses

Projected mean monthly temperatures for each GCM/SSP/year combination (n=20) were calculated in R, version 4.5.1 (R Core Team, 2025) using package ‘terra’ (Hijmans et al., 2022). Using geospatial data from ‘rnaturalearth’ package in R (Massicotte and South, 2025), and county-level census polygons from TIGER data through ‘tigris’ package in R (Walker, 2025), we constructed clean county polygons recovering coastal complexity to 20m resolution, and aligned projections to climate data. County level demographic data and projections (2000, 2030, 2050) were merged to county polygon data, and climate layers were cropped and masked to Florida. Monthly rasters were constrained to the transmission suitability temperature bounds to generate binary outcomes at the pixel level, and the number of months of suitability across the year were also calculated pixel-wise. Pixel-wise season length estimates were summarized to the county level, as mean months of suitability, and combined with county-level population estimates, to derive PAR by number of months, at baseline, 2030, and 2050, for the five GCMs, for SSP2-4.5 and SSP5-8.5 scenarios (Table 2, see Supplemental Data 1 for extended county-level estimates).

**Table 2:** Population at risk (PAR) by months of suitability for transmission for A. climate baseline and Florida population in 2000, and for B. 2030 and C. 2050, for SSP2-4.5 and SSP5-8.5, for the five GCMs in Table 1, by month, and for 9 or more months for future scenarios.

| **Year** | **2000** |
| --- | --- |
| **Model** | **Baseline** |
| *Months* |  |
| 5 | 1,434,030.00 |
| 6 | 2,716,380.00 |
| 7 | 5,544,711.00 |
| 8 | 3,954,306.00 |
| 9 | 2,332,951.00 |

**B.**
| **Year** | **2030** |  |  |  |  |
| --- | --- | --- | --- | --- | --- |
| **SSP** | **245** |  |  |  |  |
| **Model** | **HAD** | **ACC** | **CMC** | **MRI** | **MIR** |
| *Months* |  |  |  |  |  |
| 5 | 0 | 0 | 0 | 0 | 0 |
| 6 | 1,269,100 | 1,985,600 | 4,484,600 | 1,753,100 | 1,771,300 |
| 7 | 3,837,300 | 4,643,900 | 5,022,600 | 4,876,400 | 7,735,900 |
| 8 | 8,267,700 | 5,810,700 | 5,365,900 | 9,134,800 | 6,257,100 |
| 9 | 4,164,200 | 5,098,100 | 4,308,600 | 3,860,400 | 3,860,400 |
| 10 | 2,086,400 | 4,249,700 | 5,516,800 | 2,163,300 | 5,073,800 |
| 11 | 2,163,300 | 2,910,500 | 0 | 2,910,500 | 0 |
| 12 | 2,910,500 | 0 | 0 | 0 | 0 |
| 9 + | 11,324,400 | 12,258,300 | 9,825,400 | 8,934,200 | 8,934,200 |
| **SSP** | **585** |  |  |  |  |
| **Model** | **HAD** | **ACC** | **CMC** | **MRI** | **MIR** |
| *Months* |  |  |  |  |  |
| 5 | 0 | 0 | 213,000 | 0 | 0 |
| 6 | 0 | 1,686,600 | 4,718,000 | 1,686,600 | 1,967,600 |
| 7 | 4,778,100 | 3,419,800 | 5,107,800 | 4,050,700 | 5,875,500 |
| 8 | 1,851,400 | 4,400,800 | 4,834,300 | 7,821,200 | 7,908,300 |
| 9 | 9,545,800 | 8,031,100 | 4,751,600 | 3,979,800 | 3,873,300 |
| 10 | 3,449,400 | 4,162,600 | 2,163,300 | 4,162,600 | 5,073,800 |
| 11 | 2,076,200 | 2,997,600 | 2,910,500 | 2,997,600 | 0 |
| 12 | 2,997,600 | 0 | 0 | 0 | 0 |
| 9 + | 18,069,000 | 15,191,300 | 9,825,400 | 11,140,000 | 8,947,100 |

**C.**
| **Year** | **2050** |  |  |  |  |
| --- | --- | --- | --- | --- | --- |
| **SSP** | **245** |  |  |  |  |
| **Model** | **HAD** | **ACC** | **CMC** | **MRI** | **MIR** |
| *Months* |  |  |  |  |  |
| 5 | 0 | 0 | 0 | 0 | 0 |
| 6 | 0 | 1,405,700 | 1,873,700 | 1,807,300 | 1,419,700 |
| 7 | 5,445,300 | 3,513,100 | 3,964,700 | 3,834,600 | 6,226,100 |
| 8 | 2,769,500 | 2,727,000 | 1,807,400 | 5,346,000 | 3,342,100 |
| 9 | 9,984,400 | 10,553,400 | 11,367,800 | 9,333,600 | 11,134,700 |
| 10 | 4,436,100 | 4,436,100 | 1,307,900 | 2,313,800 | 2,726,500 |
| 11 | 2,213,800 | 2,213,800 | 4,527,600 | 2,213,800 | 3,216,200 |
| 12 | 3,216,200 | 3,216,200 | 3,216,200 | 3,216,200 | 0 |
| 9 + | 19,850,500 | 20,419,500 | 20,419,500 | 17,077,400 | 17,077,400 |
| **SSP** | **585** |  |  |  |  |
| **Model** | **HAD** | **ACC** | **CMC** | **MRI** | **MIR** |
| *Months* |  |  |  |  |  |
| 5 | 0 | 0 | 0 | 0 | 0 |
| 6 | 1,553,800 | 0 | 1,354,500 | 480,400 | 435,600 |
| 7 | 3,751,100 | 4,404,900 | 4,483,900 | 5,358,000 | 5,402,800 |
| 8 | 4,466,100 | 4,273,900 | 5,149,500 | 1,807,400 | 2,376,400 |
| 9 | 9,503,300 | 10,595,500 | 8,213,500 | 11,555,600 | 1,2106,700 |
| 10 | 2,848,300 | 3,361,000 | 5,647,700 | 3,433,900 | 2,313,800 |
| 11 | 2,726,500 | 2,213,800 | 3,216,200 | 2,302,800 | 2,302,800 |
| 12 | 3,216,200 | 3,216,200 | 0 | 3,127,200 | 3,127,200 |
| 9 + | 18,294,300 | 19,386,500 | 17,077,400 | 20,419,500 | 19,850,500 |

To better illustrate and visualize the impact of changing WNV transmission suitability across GCMs and projected future scenarios on the counties containing Florida’s major metropolitan centers, we plotted the months of suitability in the 10 most populous counties at baseline. The 10 counties were plotted to reflect a north to south gradient as follows: Duval (DUV), Volusia (VOL), Orange (ORA), Polk (POL), Pinellas (PIN), Hillsborough (HIL), Brevard (BRE), Palm Beach (PAL), Broward (BRO), and Miami-Dade (MIA). These counties contain many of the most densely populated metropolitan areas in Florida, including Jacksonville (Duval county), Miami (Miami-Dade county), Tampa (Hillsborough county), Orlando (Orange county), and St. Petersburg (Pinellas county).

## Results

For baseline climate, all Florida counties are suitable for WNV transmission by *Cx quinquefasciatus* for 5 or more months of the year, ranging from a 9 month season in Miami-Dade and Monroe counties, to lows of 5 months in 22 counties along the panhandle and northern border (e.g. Holmes, Jackson, Okaloosa, Escambia) (Figure 1). The PAR in 2000 at baseline climate ranges from 1.43 million (approximately 9% of the population) at risk for 5 months of the year to 2.33 million (approx. 14%) at 9 months across 2 counties, with the majority at risk in the 6-8 month range (12.22 million, approx. 73%), across 43 counties (Table 2).

**Figure 1:**
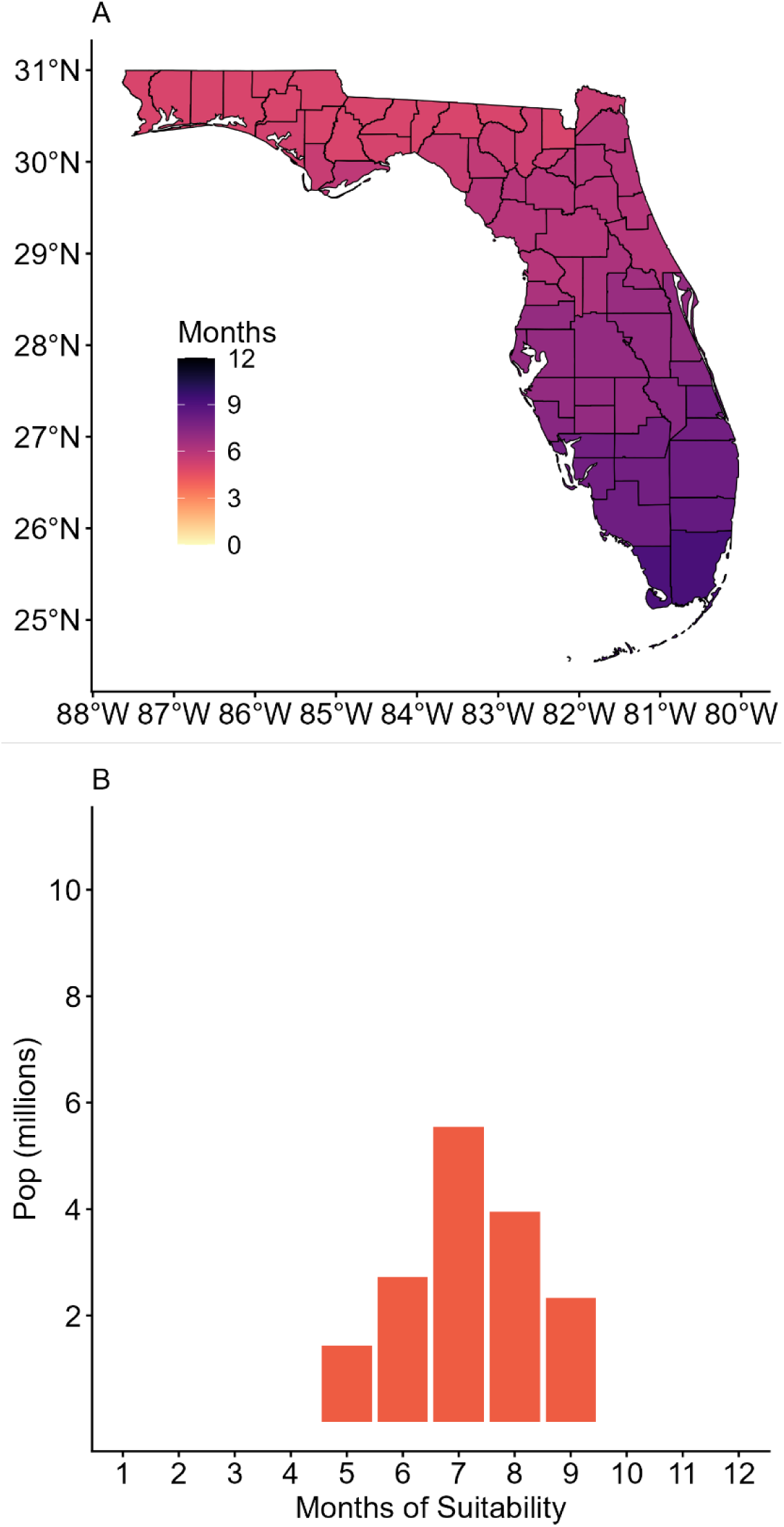
Baseline months of thermal suitability for transmission of West Nile virus by *Culex quinquefasciatus* in Florida, a. Mapped months of suitability by county; b. People at risk (PAR) by month.

Using county level population projections, we found that in 2030 and 2050 for SSP 2-4.5 and SSP 5-8.5 that every county is projected to experience 6 or more months of suitability, in all but one scenario (SSP 585, CMC, 2030). In 2030, 8.93-12.26 million people (approx. 35-48% of the population, living in 10-20 counties) under SSP2-4.5, and 8.95-18.10 million people (approx. 35-70% of the population, living in 11-26 counties) under SSP5-8.5 are projected to experience 9 or more months of suitability. In 2050, under both SSP 2-4.5 and SSP 5-8.5, 17.08-20.42 million people (approx. 58-70% of the population, living in 23-26 counties) will experience 9 or more months of suitability (Table 2, Figure 2, Supplemental Data).

**Figure 2:**
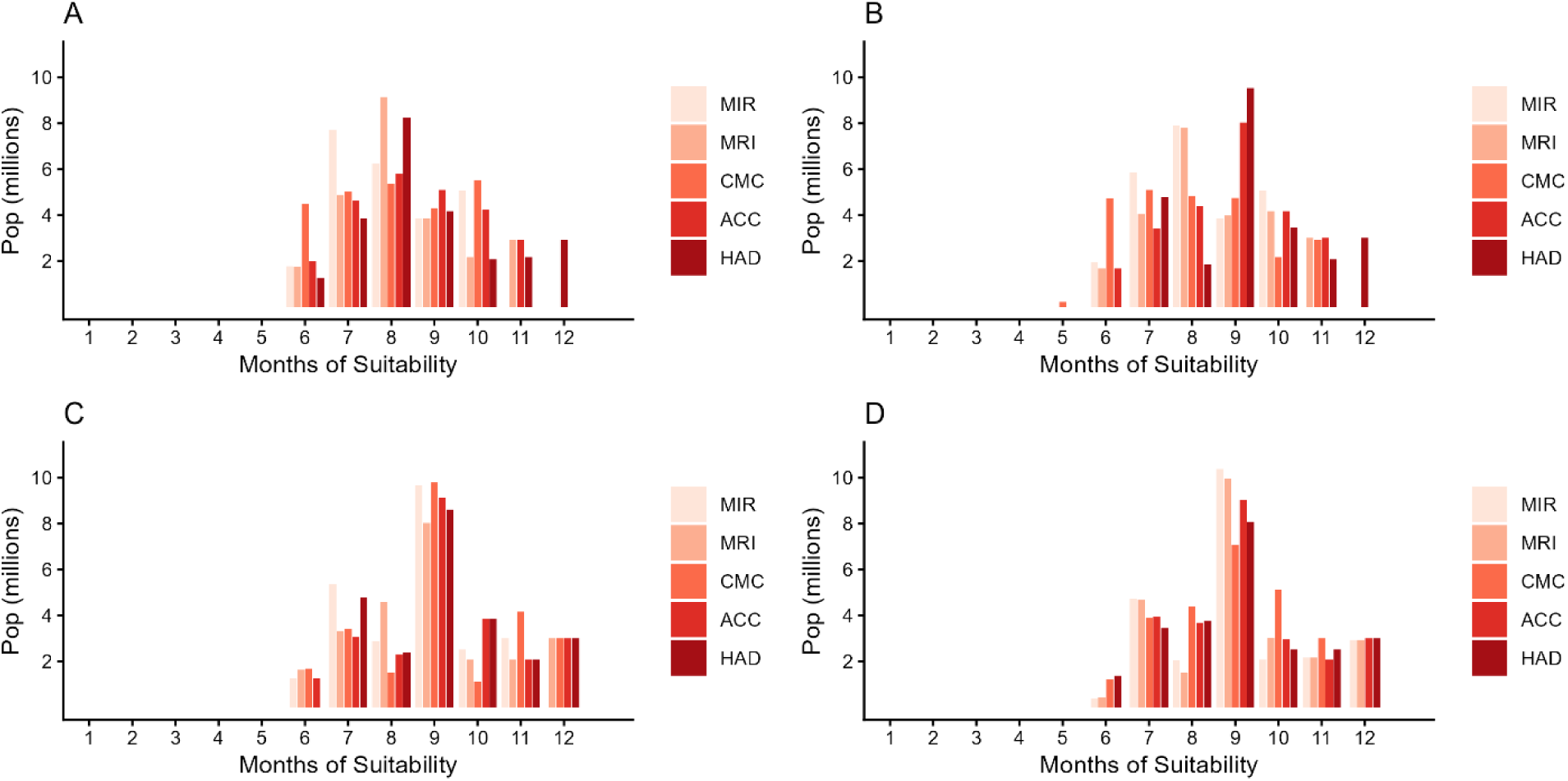
Population at risk (PAR) by months of suitability for WNV transmission in Florida, for A. 2030 and SSP2-4.5; B 2030 and SSP5-8.5; C. 2050 and SSP2-4.5; D. 2050, SSP5-8.5

The illustration of the 10 most populous counties in Florida at baseline highlights patterns in shifting WNV suitability (Figure 3). Namely, while the majority of scenarios indicate increases in the number of months with suitable conditions for transmission, ranging from 1 month to 3 months. These increases generally become more pronounced moving along the north-south gradient. For example, where Duval county in the North may gain an additional month of suitability in most scenarios, the southernmost county of Miami-Dade is projected to gain several months of suitability, resulting in year-round WNV transmission suitability (12 months) in nearly every scenario by the year 2050.

**Figure 3:**
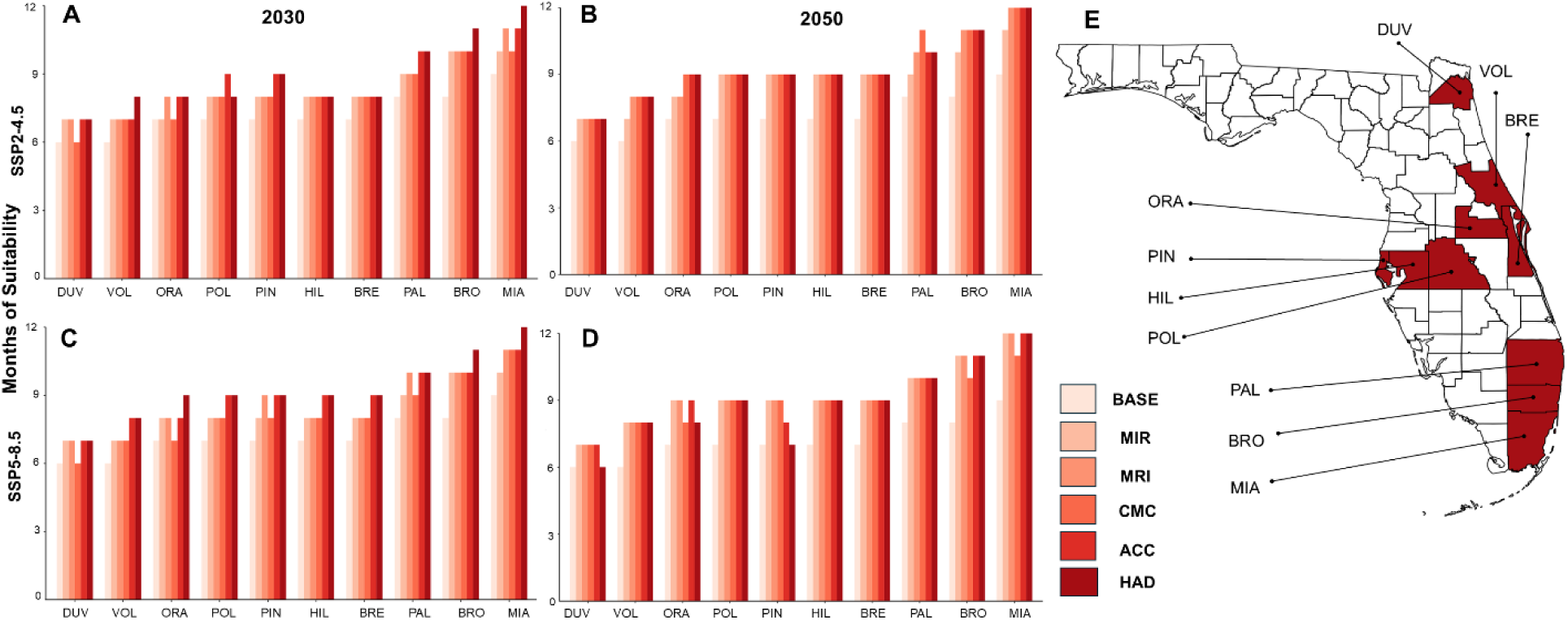
Months of WNV transmission suitability for the 10 most populous counties in Florida. Shown for baseline (BASE) and the five GCMs for the year 2030 at SSP2-4.5 (A) and SSP5-8.5 (C), and for the year 2050 at SSP2-4.5 (B) and SSP5-8.5 (D), with the counties shown on a map of Florida (E).

## Discussion

West Nile virus (WNV) is an increasingly recognized public health threat in the USA, and human cases are reported in Florida every year. Understanding shifting transmission risk in a changing environment is essential to management, both for direct intervention via vector control, and for healthcare and public awareness. In this study, at baseline climate conditions, all counties in Florida experienced 5 or more months of transmission suitable temperatures for WNV. This indicates every county needs integrated WNV surveillance and mitigation practices, including testing capacity for WNV in sentinel chickens, targeted mosquito trapping for *Cx. quiquefasciatus*, and mitigation in response to nuisance mosquito calls. These surveillance and vector control activities should be accompanied by publicly disseminated information and seasonal advisories to raise awareness of potential WNV exposure.

Recent work has demonstrated the value of the comprehensive sentinel chicken network in Florida for understanding seasonality and environmental drivers of WNV seroconversion (in the chickens) over the past 20 years (Baecher et al., 2025), yet also reveals coverage gaps remaining in some counties. The authors found that climate drivers were important at monthly and seasonal (defined as December-June) scales, with lagged minimum and maximum temperatures promoting and impeding seroconversion, respectively (Baecher et al., 2025), suggesting the important role of temperature bounds on transmissibility of WNV in Florida. As transmission season gets longer in coming years, across much of the state, so will the burden on current vector control services, including surveillance activities. Counties with substantial shifts in the number of transmission suitable months (such as Broward, Miami-Dade, and Monroe) will experience increased needs for service capacity, resources, and trained personnel. Conceptually, there may also be a need to shift the focus of services from management of nuisance mosquitoes to public health intervention, particularly in counties where extended seasonal risk coincides with WNV outbreak years.

To explore the range of projected outcomes from multiple climate models, we presented five general circulation models (GCMs) across a range of sensitivities. While every model and scenario projection show increasing numbers of people at risk in longer seasonal exposure, a minimum increase from 2.33 million people in 2 counties, or approximately 14% of the population of Florida at the time, to 8.93 million people in 10 counties (approx. 35% of the population) experiencing 9+ months of suitable temperatures in 2030, and 17.8 million in 18 counties (approx. 58% of the population) in 2050, represents a substantial increase in potential exposures and management needs, assuming all the best case scenarios. On the upper end of projections, this jumps to 18.1 million in 26 counties (approx. 70% of the population) exposed to 9 or more months of suitable temperatures in 2030, with an increase in PAR to 20.4 million in 26 counties (also approx. 70% of the projected population) in 2050. It is important to note that these years represent midpoints in the climate model projections, and we are currently within the temporal range of the first projection (2021-2040), rather than at our baseline.

Florida faces the intersection of increasing human populations and longer seasons of WNV transmission suitability in many counties. Where vector control districts may currently employ seasonal workers, for a few months of the year, for vector surveillance (e.g. trapping and identification, monitoring sentinel chicken results), and public education during ‘mosquito season’, an expanded season compounded with more coverage needs as populations increase, will mean greater and different needs. As job needs and obligations shift from seasonal, sometimes casual, employment to six or more months, or from part to full-time, this places novel constraints on budgets and necessitates additional allocations to vector control districts, which are currently managed and financed, from municipal to county, and under a variety of different budgeting models. In addition, employees themselves will become more place-bound, resulting in shifting housing needs and costs.

Expanding training for point-of-care (POC) healthcare providers in the recognition and diagnostics of arboviruses may also be needed in areas increasing in risk. Currently, there is no widely available vaccine for WNV in humans, making mosquito control and mitigation the primary tools for preventing additional cases in humans. Raising awareness of potential increases in arboviral transmission for clinical providers and promoting the adoption of personal protective behaviors (e.g., wearing mosquito repellants, limiting outdoor exposure during peak mosquito activity, etc) in communities with extended risk could help reduce the health burdens associated with shifting risk.

While suitable temperatures are a necessary component of the WNV transmission cycle, increasing temperature suitability does not directly or alone act to promote increases in vector populations or WNV cases. Many environmental disturbances can promote increases in suitable vector habitat and arboviral transmission, such as habitat fragmentation (Burkett-Cadena and Vittor, 2018; Tavares et al., 2024), urban development (Wilke et al., 2018), and natural disasters (Moise et al., 2024). Further, zoonotic arboviral outbreaks are influenced by transmission in wildlife host populations, where epizootic transmission and mosquito host preferences can drive spillover to dead-end hosts (Heidecke et al., 2023). Nevertheless, the net increase in thermally suitable areas for longer portions of the year sets the stage for increased or prolonged transmission, at times when local mosquito control districts may not anticipate activity, based on historical trends. This work provides a tool for anticipating shifts in temperature mediated WNV exposure at the county level, enabling mosquito control districts to make informed adjustments to current control programs to increase future awareness and monitoring.

## Conclusion

This study provides a useful tool to visualize and explore shifting seasonal exposure to suitable temperatures for transmission of WNV within the decade (2030) and in the longer term (2050), using two projection scenarios to capture the differences in a future with and without climate mitigation. Florida is facing the perfect storm of increasing populations, urbanization and climate change, putting millions more people at risk of shifting WNV transmission suitability in coming decades. This county-level projection of shifting risk can inform resource allocation and planning for vector control and public health across the state.

## Data Availability

All data produced in the present work are available online at: https://github.com/sjryan3/wnv_FL

## Supplemental Data

Supplemental Data and descriptions can be found at https://github.com/sjryan3/wnv_FL, which contains S1.csv - the number of months of suitability at the county level at baseline climate; S2. csv - the number of months of suitability at the county level for every combination of Year, GCM and SSP.

